# The efficacy and safety of Acupoint herbal patching in treating peptic ulcer: protocol for a systematic review and meta-analysis

**DOI:** 10.1101/2024.09.21.24314138

**Authors:** He Wang, Yuxin Jiang, Qiaoyue Sun, Jinying Zhao, Qi zhang, Fuchun Wang

**Affiliations:** Department of Acupuncture and Tuina, Changchun University of Traditional Chinese Medicine, Changchun, Jilin, China

## Abstract

**Introduction:** Peptic ulcer (PU) is prone to recurrence and can have a prolonged course, significantly impacting patients’ quality of life. Clinical treatment commonly involves combating Helicobacter pylori(HP), reducing gastric acid secretion, and promoting gastric mucosal protection. Nevertheless, Western medicine often entails various adverse effects and long-term use. Consequently, numerous scholars have redirected their focus towards traditional Chinese medicine (TCM) for external treatments of PU due to its minimal toxicity, fewer side effects, and lower recurrence rates. This study aims to assess the efficacy and safety of Acupoint herbal patching (AHP) in treating PU, offering a foundation for future clinical investigations.

**Methods and analysis:** The computer will conduct a comprehensive search for relevant studies on the utilization of AHP in the management of PU from the inception of the database in various scholarly platforms including China Journal Network, Wanfang Database, Chongqing Wipo Database, China Biomedical Literature Database, PubMed, and Cochrane Library. Eligible literature will undergo meticulous scrutiny based on predefined criteria, with data extraction and quality assessment executed independently by two researchers. Meta-analysis utilizing RevMan 5.4.1 software will be employed to synthesize the collected data. The study will focus on the TCM Symptom Score Scale as the primary outcome measure, while secondary outcomes will encompass serum inflammatory factors, endoscopic findings, quality of life, recurrence rate, and adverse events. Furthermore, assessments on effectiveness, cure rate, and potential publication bias will be carried out. This investigation aims to assess the efficacy of AHP in the treatment of PU and its impact on enhancing the well-being of patients.

**Ethics and dissemination:** Since the present work constitutes a literature review, it is important to note that ethical approval is deemed unnecessary. The outcomes of this investigation are intended for dissemination in a scholarly periodical subject to peer review.

**PROSPERO registration number:** CRD42023456995

**STRENGTHS AND LIMITATIONS OF THIS STUDY:** A thorough examination of the literature will be undertaken across six electronic databases in both Chinese and English languages.

The methodology will adhere to the guidelines outlined in the Preferred Reporting Items for Systematic Reviews and Meta-Analyses (PRISMA).

Evaluation of the studies’ quality will be conducted utilizing the updated Cochrane Risk of Bias 2.0 tool.

Variations in patch locations and treatment protocols may introduce significant heterogeneity, posing challenges to the synthesis of data.

## Introduction

PU is predominantly found to manifest in the distal esophagus, proximal duodenum, and the antral region of the stomach.^[1]^ Pathological alterations, exemplified by inflammation, mucosal lesions, or necrosis within the digestive tract, occur as a consequence of exposure to diverse inflammatory agents.^[2]^ The pain associated with this condition is distinguished by its periodic and rhythmic nature, commonly presenting alongside symptoms like indigestion, acid reflux, nausea, belching, and other related manifestations. In more severe instances, potential complications may arise, including but not limited to bleeding, perforation, obstruction, and the development of cancer.^[3]^ Significant implications on the quality of life experienced by individuals. Patients with PU tend to be between 25 and 64 years of age, and the incidence of PU is positively correlated with age. ^[1]^Based on a comprehensive examination of data from the United States, United Kingdom, and Europe, it has been established that the incidence rate of PU is 1-2/1000 individuals annually.^[4-[6]^ HP stands as the principal etiological agent of PU, displaying no age restriction in disease onset, with a notable association observed between advancing age and the prevalence of PU.^[7]^ PU is more prone to be elicited by the presence of HP infection and the utilization of NSAID drugs.^[8]^ Helicobacter pylori, a gram-negative bacterium, establishes colonization within the gastric mucosa, rendering individuals susceptible to gastritis and potential PU disease post-infection, with a subsequent risk of developing gastric cancer^.[9-10]^ Nonsteroidal anti-inflammatory drugs (NSAIDs), such as aspirin, are frequently utilized as pharmaceutical agents known to elevate the likelihood of gastrointestinal adverse reactions, notably PU. Empirical evidence indicates that individuals consuming non-aspirin NSAIDs exhibit a greater comparative susceptibility to the manifestation of symptomatic ulcers in contrast to those administered aspirin.^[11]^PU formation predominantly correlates with HP infection and NSAID consumption, yet it is also linked to gastrinomas such as Zollinger-Ellison syndrome, smoking, and heightened gastric acid secretion.^[12-14]^ Proton pump inhibitors (PPIs) have significantly revolutionized the management of PU, serving as the cornerstone of treatment for peptic ulcer-induced gastrointestinal bleeding. The length of PPI therapy post-peptic ulcer diagnosis varies depending on the specific ulcer etiology, location, and any accompanying complications. The primary objective of PPI treatment is to facilitate ulcer recovery by inhibiting acid production, concurrently addressing the root cause of the ulcers. Targeting HP eradication stands as the fundamental therapeutic target, with patients experiencing NSAID-induced ulcers advised to steer clear of aggravating agents.^[15]^ It is advised that individuals diagnosed with PU who necessitate continual NSAID treatment should persist with PPI combination therapy throughout their medical regimen.^[16-18]^ There have been changes made to corresponding prescription practices due to safety concerns about long-term PPI use, which has also caused discomfort among patients.^[19–20]^ The persistent utilization of PPI leads to the development of gastric hypochlorhydria and hypergastrinemia, potentially impeding the absorption of essential nutrients like calcium, iron, magnesium, and vitamin B12. This condition could also increase susceptibility to infections, as evidenced in relevant scholarly literature sources^.[21-22]^ Research has indicated that the utilization of PPI could potentially play a role in the onset of community-acquired pneumonia and the progression of chronic kidney disease, as evidenced by recent studies^.[23-25]^ The effectiveness of Western medicine in treating PU is well-established; however, a notable issue lies in the high rate of recurrence following cessation of these drugs. Notably, the use of antibiotic-based Western medicine is associated with a higher incidence of adverse effects, contributing to the rise of drug resistance and challenges in managing relapses^.[26]^ Consequently, apprehensions regarding the side effects and efficacy of conventional treatment methods have prompted numerous patients to explore complementary and alternative therapies.

In the realm of traditional Chinese medicine, the ailment known as PU frequently falls under the classifications of “stomach pain” and “plumpness.” This approach offers notable benefits for individuals afflicted with PU, as it facilitates enhancements in their quality of life by fostering mucosal regeneration and ulcer convalescence. By employing comprehensive diagnostic and therapeutic modalities, including etiological scrutiny, Chinese medicine demonstrates efficacy in ameliorating PU. Notably, AHP emerges as a particularly efficacious treatment modality for PU, distinguished by its protracted stimulative effects and the synergistic fusion of herbal remedies with acupuncture points.AHP is commonly employed in clinical settings due to its recognized benefits of environmentally friendly safety measures and straightforward application, catering to a spectrum of conditions encompassing internal ailments and pain-related disorders. These conditions range from diabetes and its associated complications,^[27]^ insomnia,^[28]^ dysmenorrhea,^[29]^ to cervical spondylosis.^[30]^

The contemporary scenario reveals an increasing acknowledgment of the benefits associated with AHP in the treatment of diverse ailments. Nevertheless, there is a notable absence of systematic assessments focusing on its utilization in the context of PU. In light of this deficiency, a thorough evaluation is undertaken by consolidating relevant clinical randomized controlled trial (RCT) studies concerning the use of AHP for PU. The primary objective of this meta-analysis is to evaluate the effectiveness and safety of AHP in addressing PU, with the ultimate goal of providing a fundamental basis for its practical implementation in the management of this condition.

## METHODS

### Study registration

The study has been formally recorded in PROSPERO under the reference number CRD42023456995 and is detailed in alignment with the guidelines outlined by the Preferred Reporting Items for Systematic Reviews and Meta-Analyses (PRISMA)^.[31]^

### Eligible criteria

#### Study designs

Randomized controlled trials assessing the effectiveness and safety of AHP, both as a standalone treatment and in combination with Chinese or Western medicine, in the management of PU will be the focal point of this review. Following an extensive search, the selected literature predominantly comprises Chinese publications without any restriction regarding publication date. Experiments involving animals, non-randomized controlled trials, literature reviews, and duplicated studies will be excluded from consideration in this review.

#### Participants

Subjects meeting clear diagnostic criteria for PU will be included in this study without limitations on gender, disease duration, or severity.

#### Interventions

The sole authorized experimental intervention sanctioned is AHP, which could be administered independently or in conjunction with a comparative group receiving either Chinese or Western medicine treatments. To reduce variability in clinical trials, the utilization of herbal-based AHP is constrained. Discrepancies in the timing and frequency of application, as well as the incorporation of additional mediums, will not be considered.

#### Comparators

Control interventions will consist of traditional Western pharmacological treatments or a combination of pharmaceuticals with AHP, including agents such as gastric mucosal protectors, proton pump inhibitors, and proprietary Chinese remedies. Emphasis will be placed on providing comprehensive documentation regarding dosage, administration techniques, and treatment duration. Comparative analyses involving various AHP interventions, AHP methods, or drug compositions will be omitted from the study.

### Types of outcomes

#### Primary outcome

The main focus of the study is to evaluate clinical effectiveness through the utilization of established measurement tools, mainly the TCM Symptom Score Scale.

#### Secondary outcomes

1. Levels of inflammatory factors in the serum such as TNF-α, IL-16, and IL-18.
2. Observations from endoscopic examinations.
3. Impact on the quality of life.
4. Rate of recurrence.
5. Unfavorable incidents.

### Search strategy

Computerized searches are conducted on the Chinese Journal Network Full Text Database (CNKI), WanFang Database (WanFang), Chongqing Vip Database (VIP), Chinese Biomedical Literature Database (SinoMed), American Medical Abstracts Database (Pubmed), and the Cochrane Library to identify clinically relevant randomized controlled studies on the efficacy of AHP in treating PU. Database searches will be carried out by employing a blend of medical search headings and free words, encompassing the disease name (e.g., PU), intervention (e.g., AHP), and study design (randomized clinical trial). A comprehensive approach will be adopted by exploring all potential combinations of search terms to ensure the inclusiveness of the studies considered. All publications related to the utilization of AHP in the treatment of PU are to be gathered from the inception of the library’s collection. The search methodology involves utilizing Chinese search terms such as “ acupoint plaster “, “ apply externally “, “apply”, “tian moxibustion”, “peptic ulcer”, “gastric ulcer”, “duodenal ulcer”, among others; and English search terms including “Acupoint Application”, “Acupoint Sticker”, “Herbal Patch”, “Peptic Ulcers “, “Gastric Ulcer”, “Duodenal Ulcer”, etc. The identification of relevant studies will be conducted through manual screening of all cited references to pinpoint potential eligible trials. The specific trials of interest are itemized in Table 1.

**Table 1.** Search strategy in each database.

| No | Search terms |
| --- | --- |
| #1 | Peptic Ulcers [MeSH Terms] |
| #2 | Gastric Ulcer [Title/Abstract] OR Marginal Ulcer [Title/Abstract] OR Duodenal Ulcer[Title/Abstract] |
| #3 | #1OR#2 |
| #4 | Acupoint herbal patching [MeSH Terms] |
| #5 | Acupoint Application[Title/Abstract] OR Acupoint Sticker[Title/Abstract] OR Herbal Patch[Title/Abstract] |
| #6 | #4OR#5 |
| #7 | Randomized Controlled Trial [Publication Type] |
| #8 | Randomized Controlled Trial [Title/Abstract] OR Randomized [Title/Abstract]OR Randomly [Title/Abstract] |
| #9 | #7 OR #8 |
| #10 | #3 AND #6 AND #9 |

### Study selection

Literature screening and data extraction are conducted by a team of two researchers, with a thorough assessment of the quality of the selected materials. The assessment for Cochrane risk of bias in the included literature is independently carried out by the two researchers as well^,[32]^ with any discrepancies in results resolved through consensus. Utilizing NoteExpress and Excel software, a comprehensive database is established for the literature, facilitating the extraction and documentation of relevant information. The visual representation of the study selection process can be found in Figure 1.

**Figure 1.**
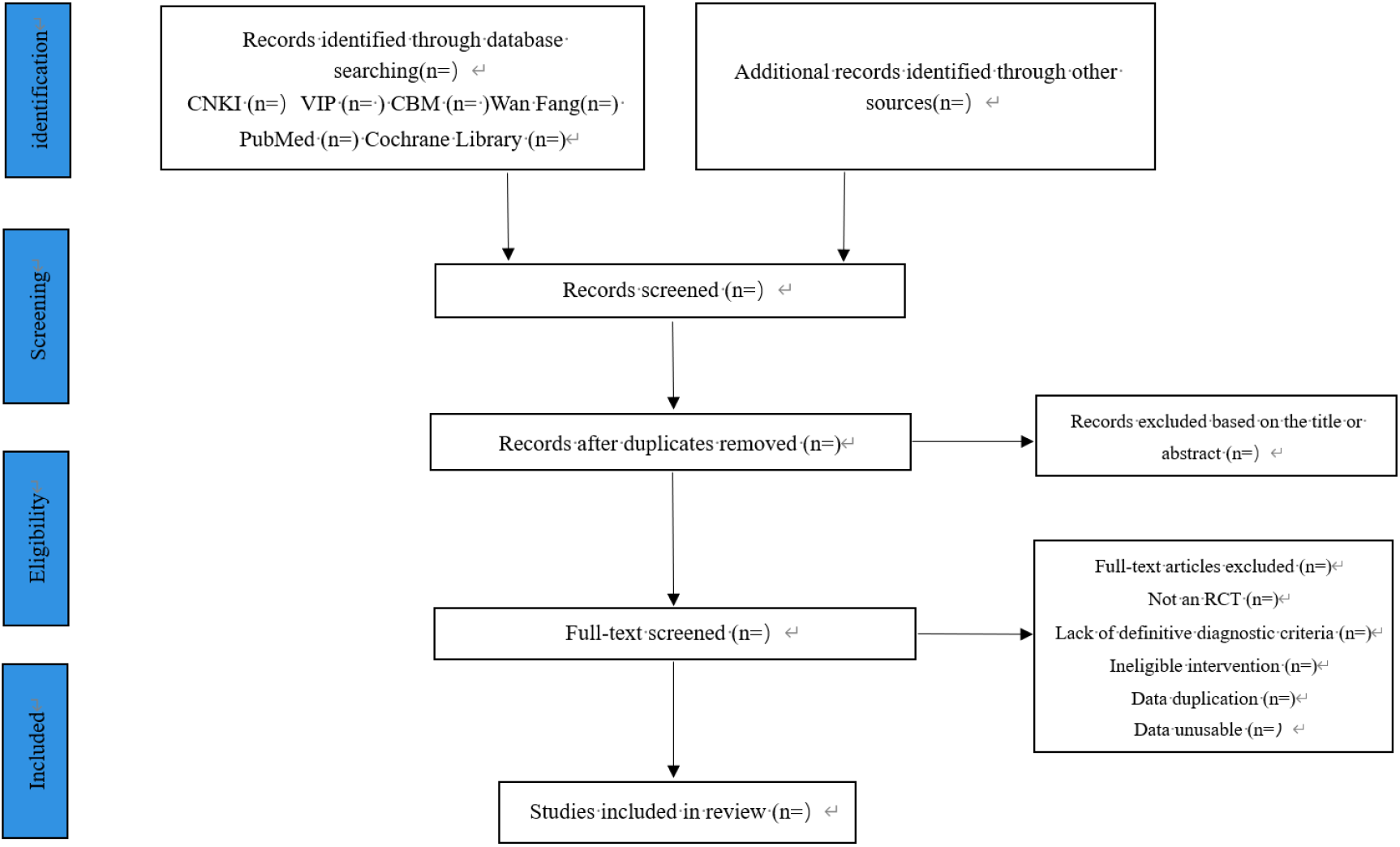
Flow diagram of the study selection process. RCT, randomized controlled trial.

### Data extraction and management

Two reviewers, identified as HW and YXJ, will be responsible for extracting various key information from the eligible studies. This includes study details such as first author, country, year of publication, language, journal, and article title; basic participant information like sample size, diagnostic criteria, mean age, gender distribution, and duration of UC; intervention specifics covering type, frequency, and duration of AHP; details regarding the control group such as drug name, dose, frequency, and treatment duration; methodological characteristics encompassing study design, randomization, allocation, and blinding; as well as outcomes, including primary and secondary outcomes. Should there be inadequate or unclear data, the corresponding author will be contacted for supplementary information. Any discrepancies that arise will be arbitrated by a third independent investigator identified as QZ.

### Assessment for risk of bias

Two independent researchers, identified as HW and YXJ, will assess the methodological quality using the ROB 2.0 tool in the context of an RCT, drawing on the Cochrane Risk of Bias 2.0 framework^.[33]^ Evaluation of the studies will involve rating the ROB across five key domains: (1) randomization process, (2) adherence to the intervention protocol, (3) handling of missing data, (4) outcome measures, and (5) selection of reported outcomes. Detailed scrutiny of each domain within individual studies will be undertaken to ascertain the level of bias present - whether it is low, high, or unclear. In cases where information is lacking or requires clarification, communication with the original study authors will be initiated. Studies with low bias across all domains will be classified as having an overall low risk of bias, while those showing bias in any domain will be categorized as high risk.^[34]^ A graphical representation of the bias summary will be provided, and any discrepancies between the two evaluators will be resolved through consultation with a third expert, denoted as QZ.

### Data analysis

#### Data synthesis

Two researchers (HW and YXJ) conduct a Meta-analysis using Review Manager 5.4.1 software, creating forest plots and funnel plots for assessing heterogeneity and detecting publication bias. Risk ratio (RR) with 95% Confidence Interval (CI) is used to express count data. Heterogeneity is assessed using the I^2^ test, with a threshold of P>0.1 and I^2^<50% indicating the use of a fixed effect model, otherwise, a random effect model is applied. Sensitivity analysis is used for large heterogeneity to ensure result stability. Descriptive analysis is conducted when the source of heterogeneity is unclear. Review Manager 5.4.1 is also utilized for generating funnel plots to investigate publication bias^.[35-36]^ Any disagreements between the researchers would be resolved by consulting a third independent investigator (QZ).

#### Assessment of heterogeneity

Assessment of statistical heterogeneity between included trials depends on I^2^. A range of I^2^ statistical values from 0% to 100% quantifies different heterogeneities.^[37]^ Negligible heterogeneity is expressed as I^2^ < 25%, mild heterogeneity as 25% ≤ I^2^ < 50%, moderate heterogeneity as 50% ≤ I^2^ < 75%, and high heterogeneity as 75% ≤ I^2^ < 100%.^[38]^

#### Subgroup analysis and sensitivity analysis

Subgroup analyses are intended to investigate potential sources of heterogeneity in cases where notable heterogeneity is present and data are deemed adequate. These analyses will be conducted by considering various characteristics of the studies included, such as the selection of distinct AHP, the application duration, medication frequency and duration, treatment duration, and disease severity. If subgroup analyses fail to elucidate the sources of heterogeneity, sensitivity analyses will be carried out or alternative statistical models will be employed to evaluate the reliability of the aggregated outcomes. Moreover, the impact of sample size, study design, and methodological quality will be carefully scrutinized.^[39]^

#### Assessment of publication bias

If the number of Randomized Controlled Trials (RCTs) exceeds ten in the context of a Meta-analysis, funnel plots will be employed to evaluate potential publication bias. In cases where asymmetry is detected within the plots, quantitative assessment will be conducted using Egger regression tests as outlined in the reference.^[40]^

#### Grading the quality of evidence

The GRADE system will be used to assess the certainty of evidence.^[41]^ The assessment of evidence quality will be conducted utilizing the Grading of Recommendations Assessment, Development, and Evaluation (GRADE) framework as outlined by references^.[42-43]^ The outcomes will be evaluated and categorized as “very low,” “low,” “moderate,” or “high” based on the GRADE rating system.

#### Patient and public involvement

No patient participation.

#### Ethics and dissemination

Ethical clearance is deemed unnecessary for this study. The findings obtained will be forwarded for consideration in reputable, peer-reviewed academic publications.

## DISCUSSION

PU impacts an estimated 5-10% of the world’s populace, exhibiting a ubiquitous presence across all regions and demographic groups, manifesting at any stage of life, and demonstrating an annual occurrence rate of around 0.1-0.3%^.[44-45]^ Approximately one-tenth of the population has experienced PU, as indicated by findings from the Chinese Population Epidemiologic Survey. The study revealed an annual occurrence rate of PU in China at approximately 0.84%, affecting a notable percentage ranging from 10.3% to 32.6% of individuals undergoing gastroscopy within the nation.^[46]^ PU presents a significant morbidity risk and is associated with various severe complications including upper gastrointestinal bleeding, pyloric obstruction, and perforation, which can pose life-threatening situations. Approximately 10% of patients necessitate surgical intervention according to data. Through extensive research into the causes and development of PU, the widespread utilization of diverse screening and diagnostic methodologies, the identification and eradication of HP, and the swift advancement and broad implementation of PU medications like PPI, the morbidity rate of PU has shown a consistent decline and a reduction in the occurrence of critical complications.^[47]^ The typical duration of a PU spans approximately 6 to 7 years, although in certain cases, patients may endure the condition for as long as 10 to 30 years. Throughout the trajectory of the disease, individuals frequently encounter episodic discomfort in the epigastric area, along with recurrent bouts of the ailment. These occurrences can potentially result in severe complications, imposing a significant toll on both the physical and mental well-being of the patient. This, in turn, leads to a decline in quality of life, disruptions in occupational functioning, increased financial burdens, and negative repercussions on societal productivity, as highlighted in the referenced source^.[48]^

The efficacy of AHP in the treatment of PU lies in its ability to ameliorate inflammatory factors, oxidative stress factors, gastrointestinal hormones, and regulate gastrointestinal function. Studies have suggested^[49]^ that AHP treatment exhibits comparable outcomes to AHP, including the inhibition of gastric acid secretion, facilitation of ulcer healing,^[50]^ mitigation of gastrointestinal adverse effects, enhancement of HP eradication rates, reduction of serum PGI and PGII levels, and lowering of recurrence rates^.[51]^

However, the advantages of AHP for PU have not been supported by a high level of evidence, and the potential value of its clinical application has not been fully revealed. Therefore, a Meta-analysis of existing RCTs is necessary. However, this study still has some limitations. The literature included in this study is exclusively in Chinese, leading to a potential language bias. Furthermore, the study is limited by a scarcity of high-quality literature and a lack of multicenter and large-sample studies. There is also inconsistency in the symptom scoring criteria used across various studies, hindering a comprehensive analysis. It is imperative to establish a standardized symptom-scoring system. Additionally, due to constraints within the literature reviewed, the study did not assess the long-term effects of AHP, which impacts the thorough evaluation of clinical efficacy.Hence, the current investigation is initiated to juxtapose the effectiveness and safety of AHP as a standalone treatment against its combination therapy counterpart in addressing PU. The outcome of our research endeavors to furnish healthcare practitioners, guideline formulators, and medical authorities with empirical insights regarding external Chinese medicinal modalities. This inquiry aims to bridge the lacuna in existing evidence concerning PU treatment utilizing AHP to a significant degree, thereby proffering a foundation for evidence-grounded clinical decisions. Moreover, this study endeavors to shape future research methodologies and steer novel trials to address the prevailing evidence deficit.

## Data Availability

All relevant data from this study will be made available upon study completion.

## Contributors

Conception or design of the study: HW and YXI. Data collection, data analysis and interpretation: HW, YXJ and QYS. Writing the manuscript: HW.Critical revision of the paper: FCW,JYZ and QZ. Approval of the final draft: FCW.

## Patient and public involvement

Patients and/or the public were not involved in the design, or conduct, or reporting, or dissemination plans of this research.

## Patient consent for publication

Not applicable.

## Provenance and peer review

Not commissioned; externally peer reviewed.

## Supplemental material

This content has been supplied by the author(s). It has not been vetted by BMJ Publishing Group Limited (BMJ) and may not have been peer-reviewed. Any opinions or recommendations discussed are solely those of the author(S) and are not endorsed by BMJ. BMJ disclaims all liability and responsibility arising from any reliance placed on the content. Where the content includes any translated material, BMJ does not warrant the accuracy and reliability of the translations (including but not limited to local regulations, clinical guidelines, terminology, drug names and drug dosages), and is not responsible for any error and/or omissions arising from translation and adaptation or otherwise.

## Open access

This is an open-access article distributed in accordance with the Creative Commons Attribution Non Commercial (CC BY-NC 4.0) license, which permits others to distribute, remix, adapt, build upon this work non-commercially, and license their derivative works on different terms, provided the original work is properly cited, appropriate credit is given, any changes made indicated, and the use is non-commercial. See: http://creativecommons.org/licenses/by-nc/4.0/.

